# Alteration of plasma von Willebrand factor in the treatment of retinal vein occlusion with cystoid macular edema

**DOI:** 10.1101/2022.02.18.22271170

**Authors:** Hiromasa Hirai, Mariko Yamashita, Masanori Matsumoto, Takeyuki Nishiyama, Daishi Wada, Naoko Okabe, Yutaro Mizusawa, Hironobu Jimura, Tetsuo Ueda, Nahoko Ogata

**Author notes:** (ext.3433), (H.H.), (T.N.), (D.W.), (Naoko Okabe), (Y.M.), (H.J.), (U.T.). (M.Y.). (M.M.). Corresponding author (<u>Nahoko Ogata</u>).

## Abstract

Retinal vein occlusion (RVO) is a major retinal disease accompanied by venous thrombosis. Although several studies have proposed an association between venous thrombosis and von Willebrand factor (VWF), the association between RVO and VWF remains unclear. We aimed to investigate the association between RVO and VWF and the alteration of VWF levels under the anti-vascular endothelial growth factor (VEGF) treatment. We enrolled 55 patients with RVO involved cystoid macular edema. They received intravitreal injection of anti-VEGF drugs, either ranibizumab or aflibercept. We examined the clinical data, measured plasma VWF: antigen (VWF: Ag) and plasma a disintegrin and metalloproteinase with a thrombospondin type 1 motif, member 13 (ADAMTS13) activity (ADAMTS13: AC) and analyzed them to identify variabilities during the treatment. At the baseline there was no significant difference between the RVO group and age-matched controls in both VWF: Ag and ADAMTS13: AC levels but ADAMTS13:AC was significantly lower in central RVO (CRVO) than in branch RVO (BRVO) (*P* = 0.015). In BRVO, the negative correlation was found between VWF: Ag and central choroidal thickness (CCT) (r = −0.51, *P* < 0.001). In BRVO after the anti-VEGF treatment, VWF: Ag decreased significantly from 134% at the baseline to 109% at 1 day after (*P* = 0.002) and to 107% at 1 month after (*P* = 0.030). In contrast, ADAMTS13: AC showed no significant difference during this period. In BRVO at 1 month after treatment, we found a negative correlation between VWF: Ag and CCT (r = −0.47, *P* = 0.001). Our findings suggest an association between VWF and CCT in patients with BRVO, thus the measurement of VWF may be useful for evaluating the disease activity and prognosis.

## Introduction

Von Willebrand factor (VWF), a large glycoprotein synthesized and secreted from vascular endothelial cells, acts as bridging molecule which adheres platelets to the injured vessels and stabilizes coagulation factor VIII [1]. VWF is specifically cleaved by disintegrin and metalloproteinase with a thrombospondin type 1 motif, member 13 (ADAMTS13) [2]. The balance between VWF and ADAMTS13 maintains the hemostaticaly coagulative function of the whole body. The release of VWF is known to increase with endothelial cell damage and elevated plasma VWF levels are usually observed in patients with hypertension and atherosclerosis [3–5]. Karaca et al. reported an association between plasma VWF and the severity of hypertensive retinopathy [6]. In other ocular diseases, elevated plasma VWF antigen (VWF: Ag) has been reported in age-related macular degeneration (AMD), and multimeric VWF has been found in pachychoroid neovasculopathy [7–9].

Several studies have also reported that increased plasma VWF levels are associated with thrombotic complications [10–12]. Interestingly, this association includes not only atherothrombotic diseases (such as myocardial infarction or ischemic stroke) but also venous thrombosis. Edvardsen et al. recently suggested an association between VWF and venous thrombosis, such as deep vein thrombosis and pulmonary embolism [12].

Retinal vein occlusion (RVO) is a major retinal disease that causes retinal hemorrhage and severe vision loss [13,14]. Vision loss in RVO is mainly due to cystoid macular edema (CME). RVO is a multifactorial disease with several risk factors, including age, hypertension, arteriosclerosis, diabetes mellitus, dyslipidemia, high blood viscosity, and thrombosis [14,15]. RVO can be classified into two types: branch RVO (BRVO) and central RVO (CRVO). In BRVO, although partial retinal ischemia may occur, blood perfusion throughout the retina is generally maintained [16]. Alternatively, the entire retina can become severely ischemic in CRVO, which leads to severe vision loss and neovascular glaucoma [17]. Usually, CRVO has a worse prognosis than BRVO. In patients with RVO, it is important to understand the retinal circulatory status to control the disease.

Intravitreal injection of anti-vascular endothelial growth factor (VEGF) drugs has been widely used for the treatment of RVO with CME. However, CME often recurs and requires additional injections. The systemic side effects of anti-VEGF vitreous injection remain controversial. Several studies have reported thromboembolic events, including myocardial infarction and ischemic stroke [18,19].

In addition, anti-VEGF antibodies are known to enter the systemic circulation, decrease VEGF levels and affect other cytokines in the plasma [20]. We have already demonstrated that VWF: Ag levels were decreased after intravitreal aflibercept injection in patients with AMD [8].

Although RVO is accompanied by venous thrombosis, few clinical studies have examined the association between VWF and RVO and these studies have reported different results. Thus, the levels of VWF in RVO remain controversial [21–23]. Furthermore, no study has investigated whether plasma VWF is influenced by anti-VEGF vitreous injection in patients with RVO. In this study, we aimed to investigate the involvement of VWF in the pathogenesis of RVO and its variability after anti-VEGF injection.

## Methods

This prospective study was conducted at Nara Medical University Hospital, Kashihara City, Nara Prefecture, Japan, from June 2014 to March 2020. This study protocol was approved by the medical research ethics committee of Nara Medical University and followed the Declaration of Helsinki. We confirmed that all methods were performed in accordance with the relevant guidelines and regulations. All patients provided written informed consent for participation in the study. Fifty-five patients diagnosed as RVO with CME who were aged 51 to 91 years and age-matched control subjects (patients before cataract surgery) were enrolled in this study. Patients who had severe systemic complications or did not receive anti-VEGF drugs, that is selected other treatments, were excluded.

RVO was diagnosed based on ophthalmological findings, slit lamp examination, fundus examination, optical coherence tomography, wide-angle fundus photography, and fluorescein angiography. We also examined best-corrected visual acuity (BCVA, LogMAR unit), central retinal thickness (CRT), and central choroidal thickness (CCT). The pathological classification (BRVO or CRVO) was judged independently by two researchers based on the examinations. All patients received intravitreal injections of anti-VEGF drugs, either ranibizumab (Lucentis; Novartis International AG, Basel, Switzerland) or aflibercept (Eylea; Bayer HealthCare Pharmaceuticals, Berlin, Germany) at a dose of 2.0 mg/0.05 mL. The drugs were selected by the attending physicians. The intravitreal injection was approached 3.5–4.0 mm posterior to the corneal limbus.

Whole blood was collected by venipuncture of the anterior arm into a tube containing 3.8% trisodium citrate (1:9). Blood samples were collected four times: before treatment (first visit), 1 day after treatment, 1 week after treatment, and 1 month after treatment. All plasma samples were stored at −80°C and thawed at 37°C prior to examination.

We measured plasma VWF: Ag levels using sandwich enzyme-linked immunosorbent assay (ELISA), rabbit anti-human VWF polyclonal antiserum (DAKO, Glostrup, Denmark) [24], and plasma ADAMTS13 activity (ADAMTS13: AC) by chromogenic ADAMTS13 activity ELISA (Kainos, Tokyo, Japan) [25]. We defined the 100% reference value as the amount of VWF: Ag and ADAMTS13: AC in pooled normal plasma from 20 volunteers (10 men and 10 women) aged between 20 and 40 years.

All statistical analyses were performed using EZR (Saitama Medical Center, Jichi Medical University, Saitama, Japan), a graphical user interface for R (The R Foundation for Statistical Computing, Vienna, Austria) [26]. We used t-tests to compare two continuous variables (such as age) and Fisher’s exact tests to compare the proportions of categorical variables (such as sex) between the groups. We also used the Friedman and Wilcoxon signed-rank tests to compare the four continuous variables (VWF: Ag). Correlations were evaluated using Pearson’s correlation coefficient. The threshold for significance was set at *P* < 0.05.

## Results

The characteristics of the patients with RVO and controls are summarized in Table 1. In patients with RVO, the median age was 73.0 years, and 33 (60%) were male. Twenty-seven (49%) patients had hypertension. There was no significant difference in VWF: Ag and ADAMTS13: AC between the RVO group and the controls (*P* = 0.70 and *P* = 0.22, respectively). We also examined the baseline characteristics of patients with RVO classified according to RVO type, BRVO and CRVO. Both CRT and CCT were significantly thicker in CRVO than in BRVO (*P* = 0.013 and 0.045, respectively). The number of dyslipidemia cases was also significantly higher in CRVO than in BRVO (*P* = 0.004). Although VWF: Ag showed no significant difference between the two groups (*P* = 0.18), ADAMTS13:AC was significantly lower in CRVO than in BRVO (*P* = 0.015). In other categories, there were no significant differences between the two groups.

**Table 1:**
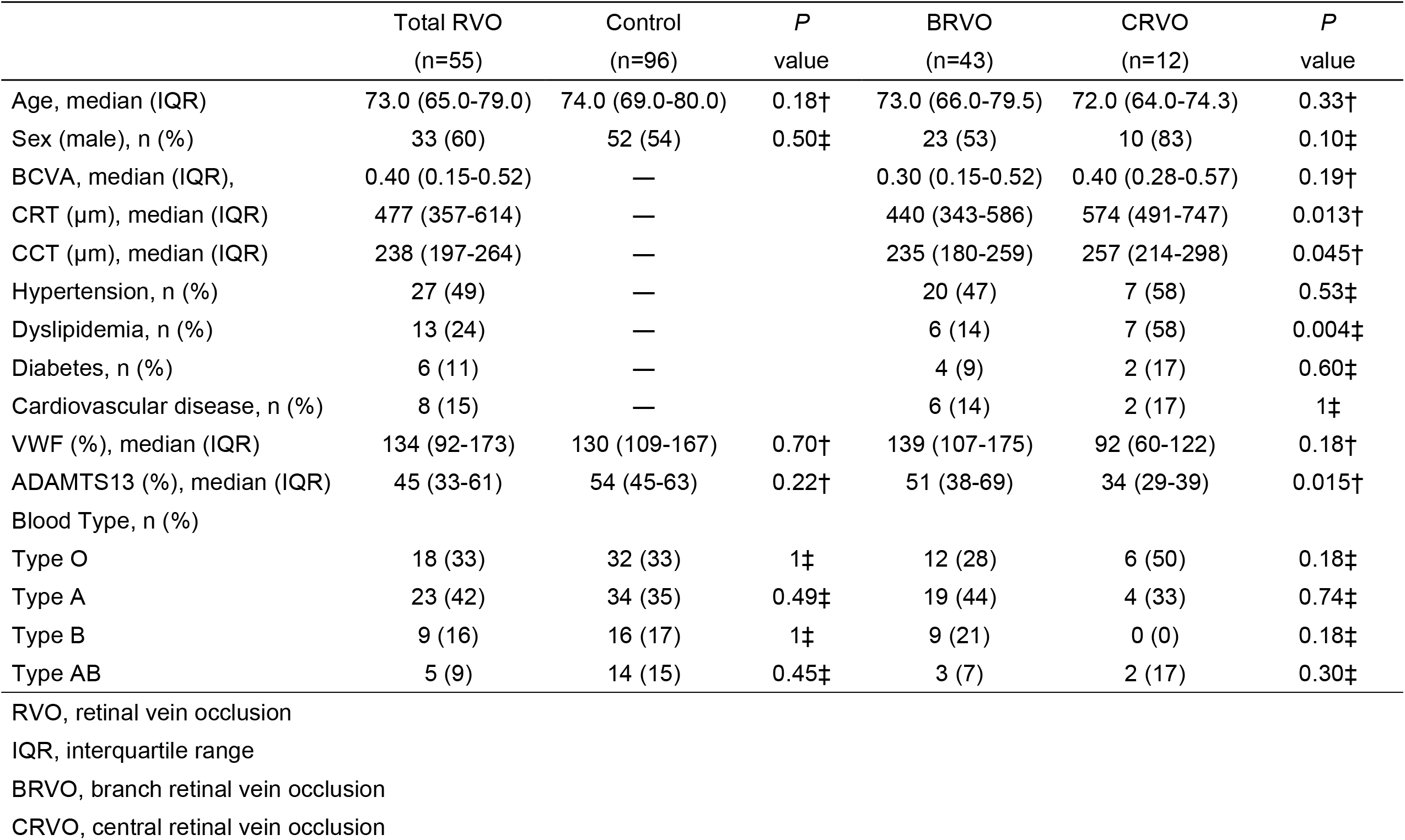
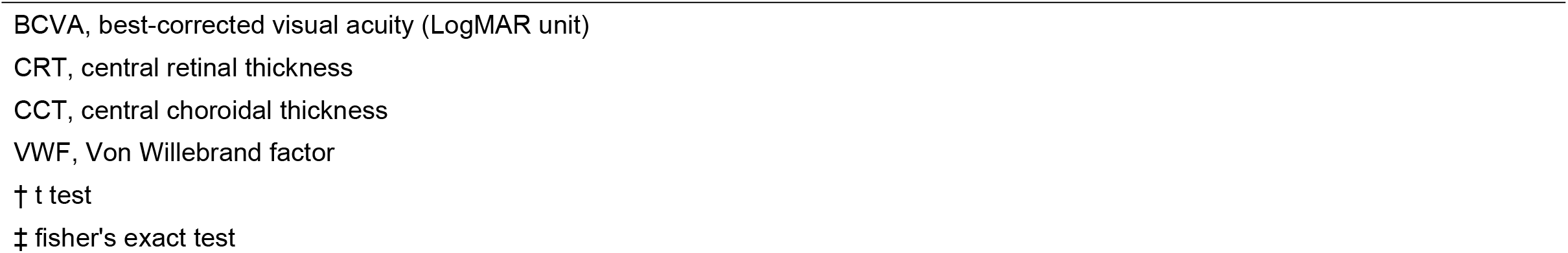
Basic characteristics of RVO patients and control.

|  | Total RVO<br>(n=55) | Control<br>(n=96) | *P*<br>value | BRVO<br>(n=43) | CRVO<br>(n=12) | *P*<br>value |
| --- | --- | --- | --- | --- | --- | --- |
| Age, median (IQR) | 73.0 (65.0-79.0) | 74.0 (69.0-80.0) | 0.18† | 73.0 (66.0-79.5) | 72.0 (64.0-74.3) | 0.33† |
| Sex (male), n (%) | 33 (60) | 52 (54) | 0.50‡ | 23 (53) | 10 (83) | 0.10‡ |
| BCVA, median (IQR), | 0.40 (0.15-0.52) | — |  | 0.30 (0.15-0.52) | 0.40 (0.28-0.57) | 0.19† |
| CRT (μm), median (IQR) | 477 (357-614) | — |  | 440 (343-586) | 574 (491-747) | 0.013† |
| CCT (μm), median (IQR) | 238 (197-264) | — |  | 235 (180-259) | 257 (214-298) | 0.045† |
| Hypertension, n (%) | 27 (49) | — |  | 20 (47) | 7 (58) | 0.53‡ |
| Dyslipidemia, n (%) | 13 (24) | — |  | 6 (14) | 7 (58) | 0.004‡ |
| Diabetes, n (%) | 6 (11) | — |  | 4 (9) | 2 (17) | 0.60‡ |
| Cardiovascular disease, n (%) | 8 (15) | — |  | 6 (14) | 2 (17) | 1‡ |
| VWF (%), median (IQR) | 134 (92-173) | 130 (109-167) | 0.70† | 139 (107-175) | 92 (60-122) | 0.18† |
| ADAMTS13 (%), median (IQR) | 45 (33-61) | 54 (45-63) | 0.22† | 51 (38-69) | 34 (29-39) | 0.015† |
| Blood Type, n (%) |  |  |  |  |  |  |
| Type O | 18 (33) | 32 (33) | 1‡ | 12 (28) | 6 (50) | 0.18‡ |
| Type A | 23 (42) | 34 (35) | 0.49‡ | 19 (44) | 4 (33) | 0.74‡ |
| Type B | 9 (16) | 16 (17) | 1‡ | 9 (21) | 0 (0) | 0.18‡ |
| Type AB | 5 (9) | 14 (15) | 0.45‡ | 3 (7) | 2 (17) | 0.30‡ |

RVO, retinal vein occlusion
IQR, interquartile range
BRVO, branch retinal vein occlusion
CRVO, central retinal vein occlusion
CRT, central retinal thickness
CCT, central choroidal thickness
VWF, Von Willebrand factor
† t test
‡ fisher's exact test

We assessed the blood type because the blood O group had lower plasma VWF: Ag levels than the other groups [27]. There was no difference in the blood type between the two groups.

Fig. 1 shows the correlation between VWF: Ag and each parameter at the baseline. In BRVO, there was a positive correlation between VWF: Ag and CRT (r = 0.30, *P* = 0.049). We also found a strong negative correlation between VWF: Ag and CCT (r = −0.51, *P* < 0.001). In CRVO, no correlations were found in any of the categories.

**Fig. 1.**
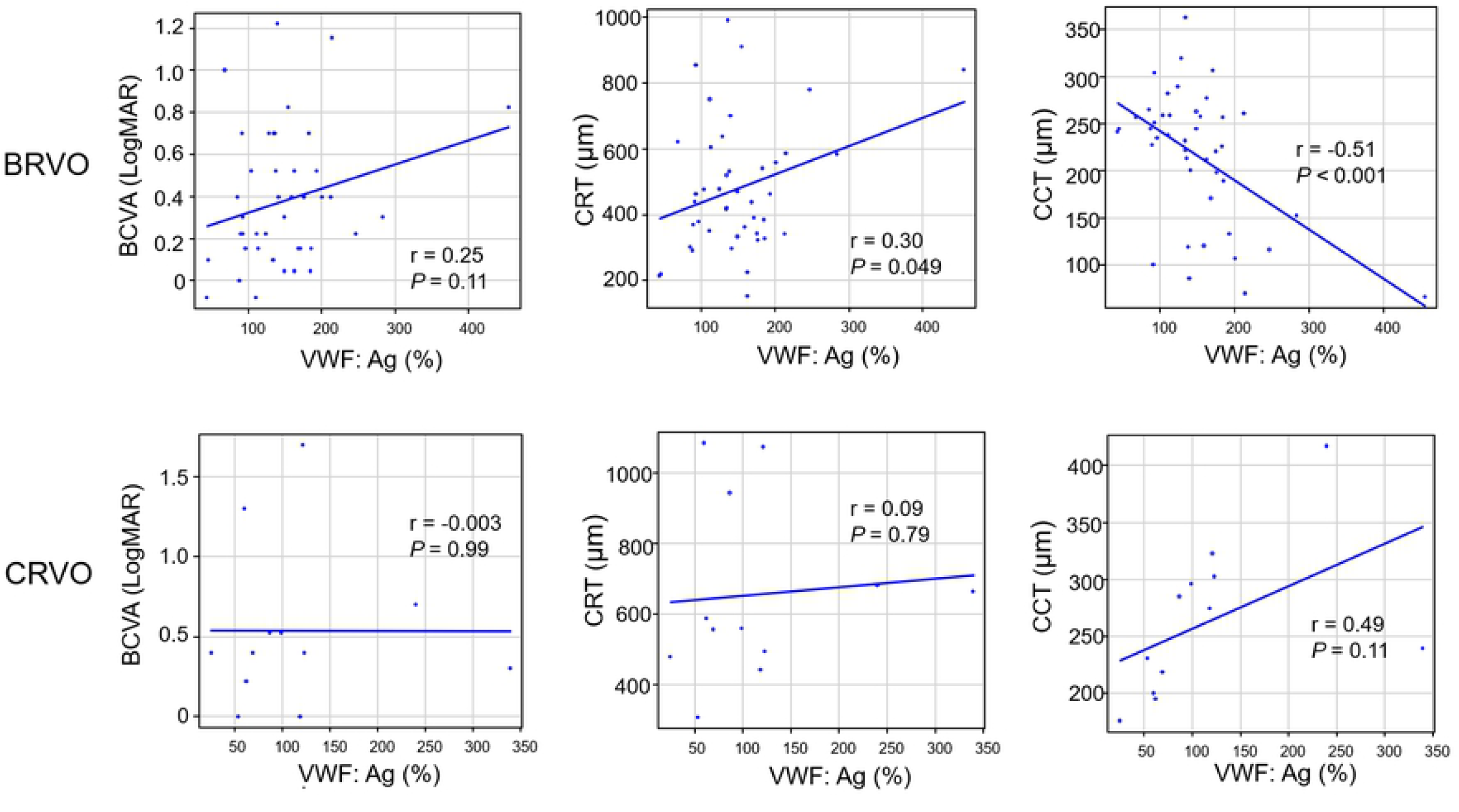
The correlation between VWF: Ag and parameters (BCVA, CRT, and CCT) at the baseline. In each graph, the horizontal axis presents the VWF: Ag (%). VWF: Ag, plasma VWF: antigen; BCVA, best-corrected visual acuity (LogMAR unit); CRT, central retinal thickness; CCT, central choroidal thickness.

Table 2 shows the basic characteristics of patients with RVO classified according to the selected anti-VEGF drugs. We confirmed that there were no significant differences between the two groups.

**Table 2:** Basic characteristics of patients classified by anti VEGF drugs.

|  | Aflibercept<br>(n=28) | Ranibizumab<br>(n=27) | *P*<br>value |
| --- | --- | --- | --- |
| Age, median (IQR) | 74.0 (66.0-79.0) | 72.0 (63.5-78.0) | 0.78† |
| Sex (male), n (%) | 15 (54) | 18 (67) | 0.41‡ |
| RVO type<br>(BRVO / CRVO) | 21 / 7 | 22 / 5 | 0.75‡ |
| BCVA, median (IQR) | 0.22 (0.14-0.52) | 0.39 (0.26-0.61) | 0.17† |
| CRT (μm), (IQR) | 456 (358-626) | 481 (371-586) | 1† |
| CCT (μm), (IQR) | 233 (199-257) | 251 (187-278) | 0.58† |
| VWF (%), median (IQR) | 118 (82-164) | 139 (111-183) | 0.54† |
| ADAMTS13 (%), median (IQR) | 40 (31-53) | 51 (34-69) | 0.67† |

IQR, interquartile range
BRVO, branch retinal vein occlusion
CRVO, central retinal vein occlusion
BCVA, best-corrected visual acuity (LogMAR unit)
CRT, central retinal thickness
CCT, central choroidal thickness
VWF, Von Willebrand factor
† t test ‡ Fisher's exact test

Table 3 shows the alterations of clinical data in patients with RVO by treatment. In total RVO, BCVA significantly improved from 0.40 at the baseline to 0.22 at 1 month after anti-VEGF treatment (*P* < 0.001). Both CRT and CCT were significantly thinner at 1 month after treatment (*P* < 0.001 and 0.001, respectively). In BRVO, BCVA also significantly improved from 0.35 at the baseline to 0.15 at 1 month after treatment (*P* < 0.001). Both CRT and CCT were significantly thinner at 1 month after treatment (*P* < 0.001 and *P* = 0.003, respectively). In CRVO, BCVA significantly improved from 0.40 at the baseline to 0.35 at 1 month after treatment (*P* = 0.004). CRT was significantly thinner at 1 month after treatment (*P* < 0.001), whereas there was no significant difference in CCT (*P* = 0.24).

**Table 3:** Alterations of clinical data in patients with RVO by treatments.

|  | Total RVO<br>(n=55) |  | *P*<br>value | BRVO<br>(n=43) |  | *P*<br>value | CRVO<br>(n=12) |  | *P*<br>value |
| --- | --- | --- | --- | --- | --- | --- | --- | --- | --- |
|  | Baseline | 1 month |  | Baseline | 1 month |  | Baseline | 1 month |  |
| BCVA, median | 0.40 | 0.22 | < 0.001† | 0.35 | 0.15 | < 0.001† | 0.40 | 0.35 | 0.040† |
| (IQR) | (0.15-0.52) | (0.05-0.40) |  | (0.15-0.52) | (0.05-0.46) |  | (0.28-0.57) | (0.21-0.40) |  |
| CRT(μm), median | 477 | 224 | < 0.001† | 440 | 222 | < 0.001† | 574 | 239 | <0.001† |
| (IQR) | (357-614) | (199-251) |  | (343-586) | (196-244) |  | (491-747) | (219-258) |  |
| CCT (μm), median | 238 | 215 | 0.001† | 235 | 213 | 0.003† | 257 | 242 | 0.24† |
| (IQR) | (197-264) | (174-270) |  | (180-259) | (160-267) |  | (214-298) | (204-290) |  |

IQR, interquartile range
BRVO, branch retinal vein occlusion
CRVO, central retinal vein occlusion
BCVA, best-corrected visual acuity (LogMAR unit)
CRT, central retinal thickness
CCT, central choroidal thickness
† paired t test

Fig. 2 shows the correlation between VWF: Ag and each parameter 1 month after anti-VEGF treatment. In BRVO, we found a negative correlation between VWF: Ag and CCT (r = −0.47, *P* = 0.001). In CRVO, no correlations were found in any of the categories.

**Fig. 2.**
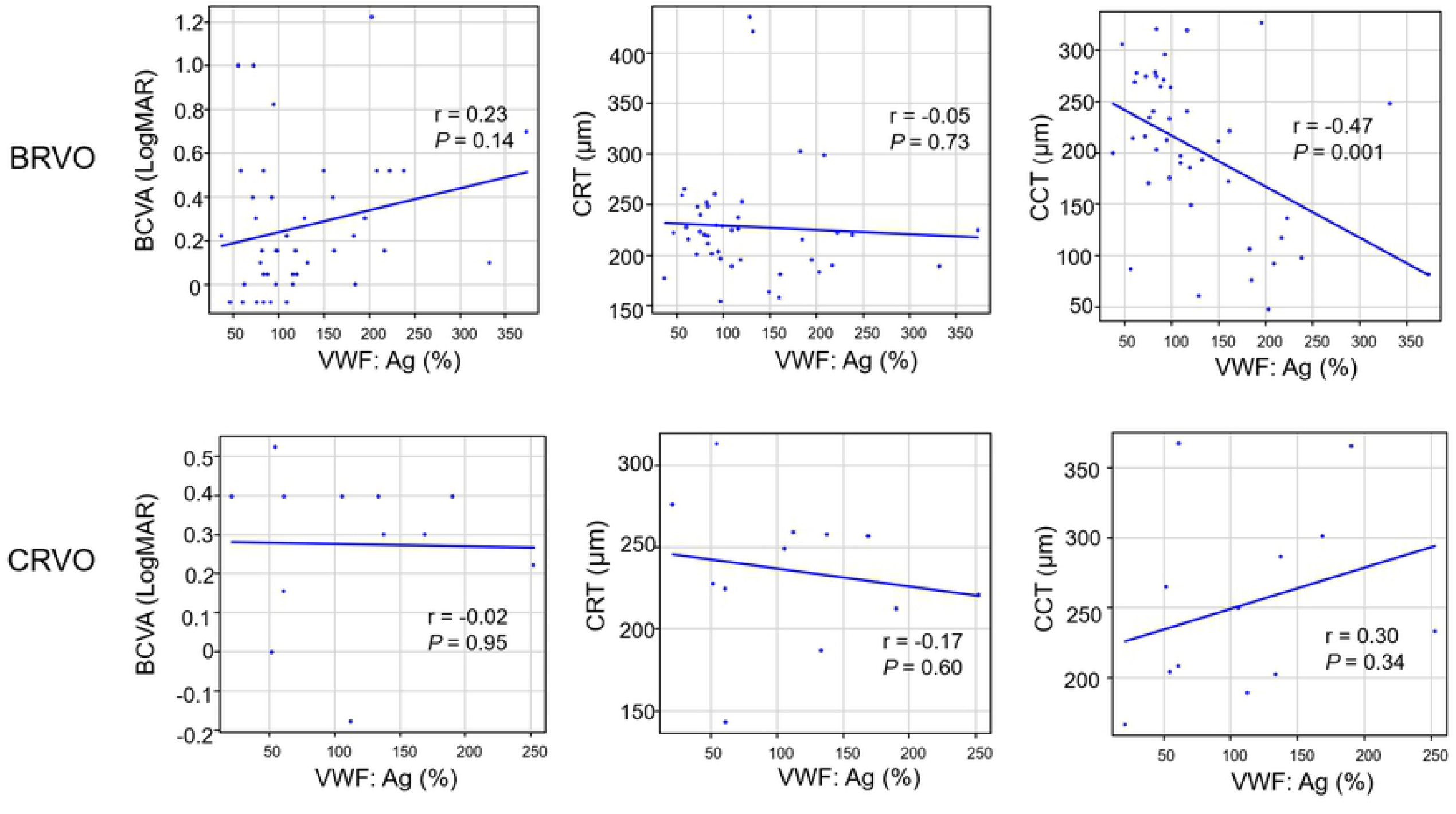
The correlation between VWF: Ag and parameters (BCVA, CRT, and CCT) 1 month after anti-VEGF treatment. In each graph, the horizontal axis presents the VWF: Ag (%). VWF: Ag, plasma VWF: antigen; BCVA, best-corrected visual acuity (LogMAR unit); CRT, central retinal thickness; CCT, central choroidal thickness.

Fig. 3 shows the time course of VWF: Ag in patients with total RVO, BRVO, and CRVO. In total RVO, VWF: Ag decreased significantly from 134% at the baseline to 104% at 1 day after treatment (*P* = 0.002). In BRVO, VWF: Ag also showed a significant reduction from 139% at baseline to 113% at 1 day after treatment (*P* = 0.002) and to 98% at 1 month after treatment (*P* = 0.030). However, in CRVO, VWF: Ag showed no significant difference during this period. ADAMTS13: AC showed no significant difference during the study period in all categories.

**Fig 3.**
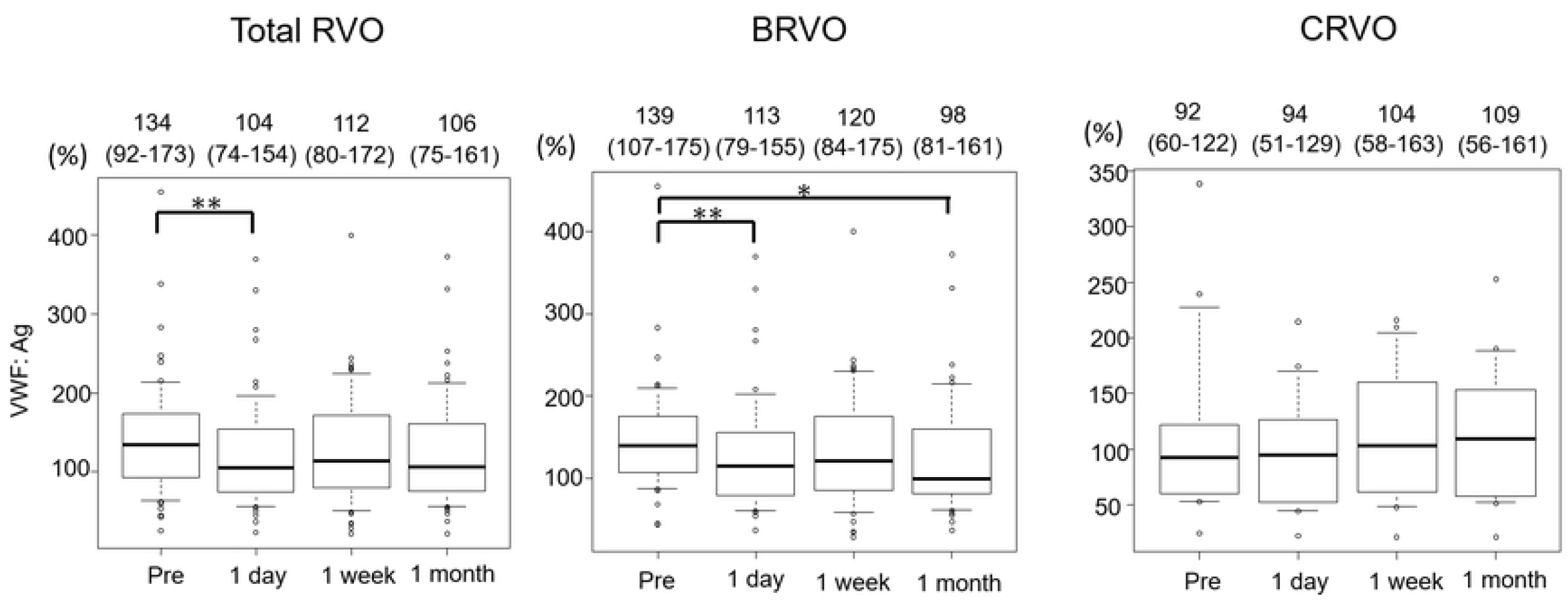
Alterations of VWF: Ag after anti-VEGF treatment (* < 0.05, ** < 0.01). Each of the values above the graph presents the median with the interquartile range.

## Discussion

This study investigated the coagulation-related factors in patients with RVO at the baseline and their alterations during the treatment.

According to our results, there was no significant difference in plasma VWF between the RVO and age-matched controls at the baseline. Several researchers have reported that plasma VWF increased not only in patients with hypertension and arteriosclerosis but also in healthy individuals with aging [28,29].

ADAMTS13:AC was significantly lower in CRVO than in BRVO at the baseline. Interestingly, the number of dyslipidemia cases was significantly higher in CRVO than in BRVO. ADAMTS13 is mainly synthesized in hepatic stellate cells [2]. Thus the liver cells in patients with dyslipidemia would be damaged by fat deposition, then production of ADAMTS13 could be decreased in patients with CRVO.

Although it was not statistically significant, the VWF levels in BRVO were higher than those in CRVO. BRVO is accompanied by a thrombus at the arteriovenous crossing point where the retinal artery and vein share an outer sheath [30,31], therefore the arterial pressure directly cause thrombosis. Alternatively, CRVO is accompanied by a thrombus in the central retinal vein where it passes through the lamina cribrosa within the optic nerve. A recent study focusing on BRVO and CRVO has shown that atherosclerosis is a more inducing factor for developing BRVO than that for CRVO [32]. High levels of plasma VWF in our study suggest the atherosclerosis in BRVO.

We also compared VWF levels and each ophthalmologic parameters of RVO and found a significant negative correlation between VWF: Ag and CCT in BRVO at the baseline. Although studies examining the association between VWF and choroidal thickness are few, Gifford et al. reported an inverse association between choroidal thickness and VWF in patients with liver cirrhosis [33]. Many previous reports demonstrated that choroidal thickness in patients with RVO decreases after intravitreal injection of anti-VEGF drugs [34]. Sakanishi et al. reported that the recurrence of CME was significantly few in cases with choroidal thinning after treatment [35]. Thus, CCT may reflect the treatment effect and prognosis in RVO. Interestingly, a significant negative correlation between VWF: Ag and CCT was still observed 1 month after anti-VEGF injection in BRVO. Therefore, the measurement of VWF: Ag during the treatment may be useful in evaluating the disease activity of BRVO. VWF: Ag also showed significant alterations under anti-VEGF treatment, especially in BRVO. Several researchers have previously proposed the association between VWF and VEGF [36,37]. VWF is secreted from endothelial-specific organelles, called Weibel palade bodies (WPBs) which widely distribute in the systemic vessels [2]. VEGF activates exocytosis of WPBs, resulting in the release of VWF [37,38] which activates VEGF-R2 signaling and may promote angiogenesis [39]. Although few studies have examined the association between anti-VEGF drugs and VWF, Pace et al. reported decreased VWF plasma levels after anti-VEGF treatment (bevacizumab) in recurrent glioma [40]. We also report the VWF reduction after anti-VEGF treatment in patients with BRVO. Thus, the alterations of VWF may reflect the therapeutic effect after anti-VEGF drug administration in BRVO.

This study has some limitations. First, this was a cross-sectional study with a relatively small sample size. A larger number of patients will be required for more accurate analyses. Second, we only assessed patients who received anti-VEGF drugs. It might be necessary to compare VWF levels in patients with and without treatment at the same time course. However, it is difficult to analyze untreated patients through frequent medical visits. Finally, we did not evaluate the long-term effects of anti-VEGF drugs for more than 1 month after intravitreal injection. A longer follow-up period is required to confirm these results.

## Conclusion

In summary, we performed prospective analyses on VWF in patients with RVO during the treatment and found unique characteristics. Our findings suggest an association between VWF and CCT in BRVO, thus the measurement of VWF may be useful for evaluating the disease activity and prognosis.

## Data Availability

The data file is available https://doi.org/10.6084/m9.figshare.19168346

## Acknowledgements

The authors thank Hiroki Tsujinaka for his useful discussions and Editage (www.editage.com) for English language editing.

